# Neural Oscillation Features of ADHD Symptoms in Children: EEG Evidence from Resting State and Oddball Task

**DOI:** 10.1101/2024.08.21.24312402

**Authors:** Siyuan Zhang, Shuting Yu, Xiaobing Cui, Xuebing Li

**Author notes:** Correspondence to: Key Laboratory of Mental Health, Institute of Psychology, Chinese Academy of Sciences, Beijing 100101, China, E-mail address (X. Li).

## Abstract

The current study examined resting-state and event-related neural oscillations associated with ADHD symptoms in children aged 6-12. 77 children were assessed using the Integrated Visual and Auditory Continuous Performance Test (IVA-CPT) and EEG during resting-state. A group of these children also completed a classic visual oddball task. Key findings include increased relative delta activity at left parietal electrodes during eyes-open and decreased relative theta activity at left posterior electrodes during eyes-closed, both associated with poor attention. Increased beta activity at right parieto-occipital electrodes during eyes-open and increased alpha activity at bilateral posterior electrodes during eyes-closed were associated with poor response control. In addition, the power of the P3 component was negatively correlated with attention across most frequency bands and conditions, except for delta power in the standard condition. Furthermore, combining multiple metrics, especially resting-state EEG oscillations, event-related oscillations, and parental ratings, provided a more robust prediction. The current study identified important brain regions and frequency bands related to ADHD symptoms, offering new insights for multi-metric prediction and clinical guidance.

## 1 Introduction

Attention Deficit/Hyperactivity Disorder (ADHD) is a common neurodevelopmental disorder characterized by problems with attention, hyperactivity, and impulsivity (American Psychiatric Association, 2013). It usually occurs in early childhood and has a global prevalence of around 5% (American Psychiatric Association, 2013). ADHD symptoms not only have academic and social consequences for children, but also increase parenting stress (Arnold et al., 2020; Galloway et al., 2019; Harpin et al., 2016; Martin et al., 2019). Furthermore, ADHD can persist into adulthood, with symptoms often evolving over time and an increased risk of developing psychiatric comorbidities such as depression, anxiety, and substance addiction (Franke et al., 2018; Katzman et al., 2017). Given the substantial burden of ADHD on individuals, families, and society, early identification and intervention are important.

Current diagnostic methods for children with ADHD primarily rely on questionnaires and interviews, which are typically provided by parents to assess their child’s behavioral performance (Danielson et al.,2018; Kieling, & Rohde, 2012). The diagnostic process is influenced by various subjective factors, especially parents’ academic expectations (Narad et al., 2015; Tahıllıoğlu et al., 2021). In clinical settings, parents are more likely to report behaviors such as lack of concentration in class and poor academic performance. Excessive concerns may lead to developmental challenges being labeled as specific medical disorders and even could potentially result in the use of stimulants to manage children’s behavior and improve academic performance (Abdelnour et al., 2022; Kazda et al., 2021). Consequently, identifying objective assessment criteria is a key goal in clinical diagnosis.

Currently, there is a growing focus on understanding the objective underlying neural mechanisms of ADHD children. A major focus is on the EEG characteristics of individuals with ADHD, which can not only aid in more accurate diagnosis but also provide guidance for non-pharmacological treatments such as neurofeedback and neuromodulation (Enriquez-Geppert et al., 2019; Lenartowicz, & Loo, 2014; Rubia et al., 2021). Although there is a long history of using EEG to assess individuals with ADHD (Jasper et al., 1983), and researchers have identified certain patterns of neural activity disorders in individuals with ADHD and have begun experimenting with specific endogenous activities as targets for neurofeedback (e.g., theta/beta, sensorimotor rhythm; Enriquez-Geppert et al., 2019; Van Doren et al., 2019), the search for definitive EEG biomarkers has had limited success.

Resting state brain oscillations and event-related potentials (ERPs) are most commonly collected to extract meaningful information. Resting state oscillations are usually analyzed by spectral analysis of average brain power across various frequency bands, revealing the state of specific frequency bands at particular spatial locations. Researchers have primarily focused on slow-wave theta and fast-wave beta activities typically recorded over frontocentral electrodes among the abnormal brain activities in ADHD (Alba et al., 2016; Heinrich et al., 2014; Hermens et al., 2005; Hale et al., 2010). The dominant view is that there is increased theta activity and deficient beta activation in individuals with ADHD, which have significant correlations with the core symptoms of ADHD (Clarke et al., 2020; Lenartowicz, & Loo, et al., 2014). Some studies have also found abnormal alpha levels, which are closely related to arousal and indicate a hypo-aroused central nervous system (Koehler et al., 2009; Lenartowicz et al., 2018). However, none of these features is sufficient as a diagnostic criterion to distinguish ADHD from neurotypical individuals.

With the development of neurofeedback techniques, the theta/beta ratio (TBR), which was previously thought to be more reliable than simply analyzing absolute and relative power, has become one of the most studied EEG measures (Lubar, 1991). Although it is the only EEG measure recognized by the US Food and Drug Administration (FDA) as an adjunctive diagnostic method in ADHD (Stein et al., 2016; http://www.fda.gov/newsevents/newsroom/pressannouncements/ucm360811.htm), and down-regulation of TBR through neurofeedback has shown some success in reducing ADHD symptoms, there is growing evidence that TBR is not robust enough to be used as a confirmatory technique for diagnosis or as a neurofeedback target (Arns et al.,2013; Gloss et al., 2016; Loo et al., 2013). On the one hand, TBR may be influenced by other frequency band components, such as the individual alpha peak frequency (Lansberg et al., 2011; Vollebregt et al., 2015). On the other hand, TBR has been found to be influenced by several moderating factors, such as age, ADHD subtype, and others (Loo et al., 2013). These findings indicate that TBR may not be a specific and reliable indicator. In summary, the resting-state EEG characteristics of ADHD are influenced by a variety of factors, and there is still no consensus on these characteristics.

Recognizing the difficulty of diagnosing ADHD based on resting-state oscillations alone, some researchers have begun to focus on the transient neural dynamics captured by ERPs. ERPs capture the temporal evolution of neural activity patterns related to attention or behavioral control and accurately reflect specific stages of processing. The P3 component (a positive voltage deflection around 300 ms over centro-parietal regions) is one of the most promising diagnostic indicators and has been associated with later attentional processing, including selective attention and response inhibition (Peisch et al., 2021; Polich, 2007; Szuromi et al., 2011). Repeated findings suggest that individuals with ADHD show a reduced P3 amplitude and prolonged latency (Kaiser et al., 2020; McPherson & Salamat, 2004; Seçen et al., 2023). In practice, stimulant medications normalize the amplitude of the P3 component, which is also associated with behavioral improvement, suggesting that targeting P3 is a promising approach for precision medicine care (Peisch et al., 2021). Some early components associated with physical processing, such as P2 and N2, have also received attention, but no consistent conclusions have been reached (Johnstone et al., 2013; Karch et al., 2010). Although empirical studies have identified some differences in ERP components between individuals with ADHD and healthy individuals, these characteristics are currently insufficient to serve as robust diagnostic criteria.

In the present study, event-related oscillations (EROs) were included in the analysis. EROs reflect the spectral characteristics within a specific time window of the task state and combine the advantages of the two common methods mentioned above. We also analyzed the resting-state EEG, exploring their relationship with children’s objective behavioral performance. Instead of focusing on a diagnosis, we used children’s behavioral performance on core ADHD symptoms as outcome variables, which is rare in previous studies. Given the heterogeneity of ADHD, this approach aims to identify both resting state and task related EEG features associated with objective performance. We did not abandon subjective assessment, as it is also important in diagnosis. Parental assessments were included to examine whether parental ratings are consistent with children’s actual performance. Additionally, we conducted exploratory regression analyses, expecting that incorporating spectral information from attentional processing may improve the accuracy of symptom prediction. Given that using the spectral characteristics of the task state rather than the resting state may serve as a better therapeutic guide for neurofeedback and neuromodulation treatments, we also expect that the current work will contribute to clinical applications.

## 2 Methods

### 2.1 Participants

77 children between the ages of 6 and 12 participated in the study. All participants were either referred to us by local primary school teachers or recruited via advertisements on WeChat. One participant was taking methylphenidate hydrochloride extended-release tablets, but stopped over 24 hours before the experiment. All participants and their parents gave written informed consent. The study was approved by the local ethics committee and followed the ethical standards of the Helsinki Declaration. As a reimbursement for their participation, children received attention training textbooks and exercise books as gifts.

### 2.2 Materials and measurements

#### 2.2.1 ADHD symptoms and intelligence assessments

ADHD symptoms were assessed using the Integrated Visual and Auditory Continuous Performance Test (IVA-CPT, Sandford & Turner, 2000). The IVA-CPT measures attention and response control in children (6 years and older), adolescents, and adults. The entire test takes approximately 20 minutes and requires participants to click the mouse only once when they see or hear “1” and not to respond when “2” appears. Upon completion of the test, the software provides summarize overall performance in response control and attention. It is currently being used as an assisted tool for the diagnosis of ADHD in several countries (Pan et al., 2007; Won et al., 2020).

Raven’s Standard Progressive Matrices (RSPM) were used to measure children’s intelligence (Raven, 1989). It is an easy-to-administer non-verbal test that is widely used to measure general cognitive ability. The entire test consists of five sets (A, B, C, D, and E) of 12 items each and takes approximately 45 minutes to complete. It is widely used and regarded as a valid indicator of general cognitive ability worldwide (Raven, 2000).

#### 2.2.2 Visual oddball task

A part of participants completed an oddball task to supplement the EEG data in the task state. For the visual oddball task, two black-and-white cartoon pictures of “apple” and “pear” were used. The “pear” was frequently presented as the standard picture (80%) and the “apple” was the target picture (20%). All of the participants were asked to press “F” quickly and accurately when “apple” was presented on the center of screen, otherwise press “J”. We required participants to response to every stimulus to assure that they attended to all items. The task contained three runs with about 30 s intermediate breaks. In each run, the “pear” was presented 80 times and the “apple” was presented 20 times (Figure 1). All the participants practiced several times before the formal experiment.

**Figure 1.**
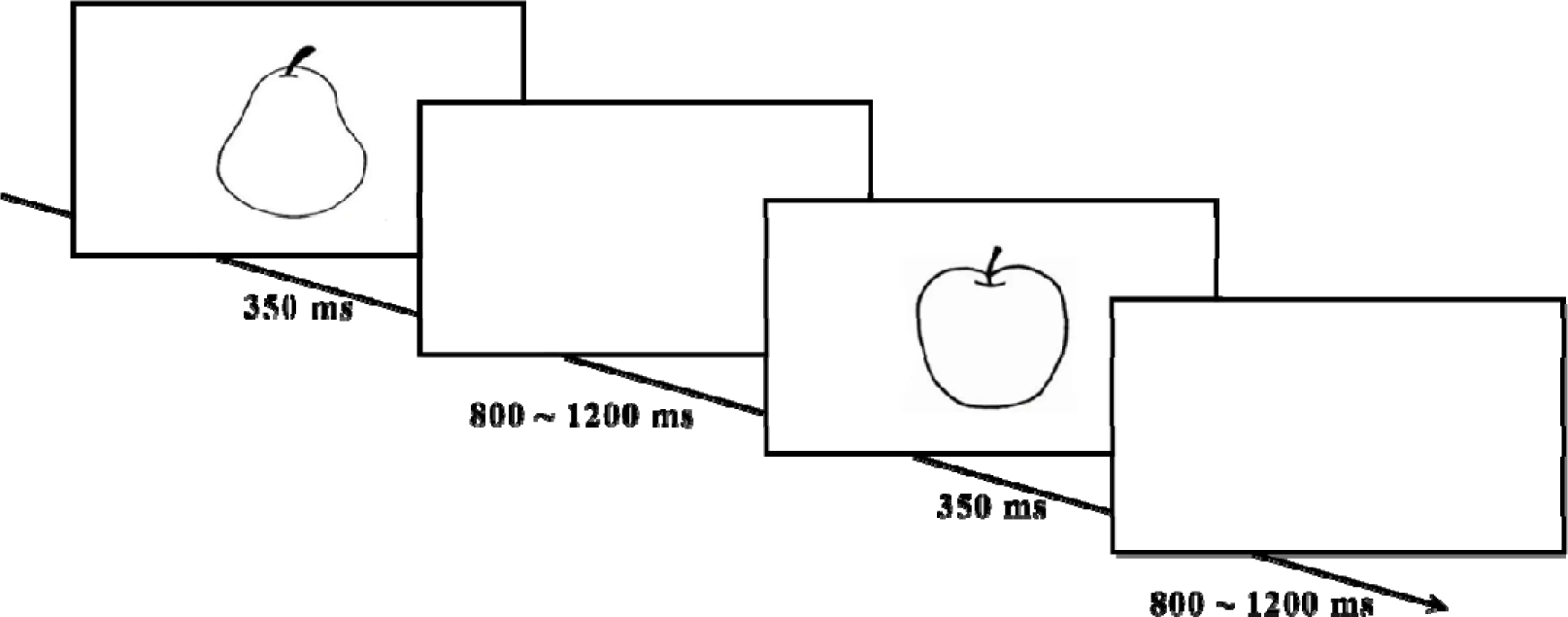
As illustrated, each stimulus was presented on a white background for 350 ms followed by a blank randomly from 800 ms to 1200 ms. The first button pressed within the blank was counted as the response for each trial. Instruction:” If you see an apple, press ‘F’, otherwise press ‘J’. The pictures are presented for a very short time, please pay attention and response as fast and accurate as possible.”

#### 2.2.3 Parental assessments

Three parent rating scales were used to assess children’s behavioral performance. ADHD Rating Scale–IV (ADHD RS-IV): Home Version is one of the most widely used ADHD scales for school-age children. It is a DSM-based scale with 18 items derived directly from the DSM-IV diagnostic criteria (DuPaul et al., 1998a; DuPaul et al., 1998b). It assesses two aspects of ADHD (inattention and hyperactivity-impulsivity) and has good discriminant validity both within the ADHD subtypes and between children with ADHD and without ADHD. The Chinese version used in the current study has proven to be a reliable and valid ADHD rating scale in China (Su et al., 2015).

The Child Behavior Checklist (CBCL) is a valid tool to evaluate children’s social skills and behavioral problems (Achenbach, 1991). It is a 118-item scale used internationally to assess children’s behavior and emotional problems and includes subscales of Attention Problems to evaluate children’s attention problems. It is available in three versions: for parents, for teachers, and as a self-report scale for older children, and the parent-report version is widely used in China (Zhang, 2005).

The Conners’ Parent Rating Scales (CPRS) is a popular research and clinical tool for obtaining parental reports of children’s behavior problems, especially for screening ADHD (Goyette et al., 1978; Deb et al., 2008). It contains 48 items in 6 dimensions, with hyperactivity and ADHD index closely related to ADHD assessment. The Chinese version of 48-items was used in the current study (Zhang, 2005).

### 2.3 EEG data acquisition and processing

EEG data were recorded using a 64-channel NeuroScan system at a sampling rate of 1 kHz. One channel placed on the left mastoid and one channel between Fpz and Fz were used as the reference and as the ground, respectively. EOG activity was monitored via two bipolar electrodes placed 1 cm at the outer canthus of each eye to measure horizontal EOG, and a separate bipolar montage was placed above and below the center of the left eye to record vertical eye movement. All electrode impedances were <10 kΩ.

EEG data were recorded during the whole experiment. At the very beginning, the resting state EEG of each participant were recorded. Eyes-open and eyes-closed data were recorded for three minutes, respectively. Participants were instructed by one experimenter to fix on a fixation or close their eyes while staying relaxed.

All data preprocessed by EEGLAB toolbox (v2021.1) in MATLAB R2020a. Raw data were filtered with a band-pass filter of 1 to 30 Hz. Resting-state data were epoched into 2-s segments, and data during the oddball task were epoched across trials and time-locked to the onset of the compound stimuli for each condition (time window: −200 – 800 ms, standard picture and target picture) with a baseline correction of 200 ms before the stimulus. Any artifactual portions of the EEG data were rejected by visual inspection. Bad channels were interpolated. For resting-state data, the middle 2-min without artifacts were retained. Synchronous or partially synchronous artifactual activity (mostly blinks) were detected on the basis of the topographical and spectral distribution and on the time series of the independent component analysis (ICA) on continuous data. Finally, data were re-referenced to the average reference.

Power spectrum analysis was conducted by Fast Fourier Transform (FFT) via MATLAB R2020a. Resting-state data were downsampled to 250Hz before FFT. Power was calculated for each frequency band, which includes delta (δ: 1-4 Hz), theta (θ: 4-8 Hz), alpha (α: 8-13 Hz), and beta (β: 13-30 Hz) for each electrode. The relative power for each frequency band was calculated by normalizing the absolute power to the total broadband power (Formula 1: *P_rel_*, relative power; *P_abs_*, absolute power).

Formula 1:

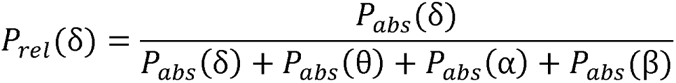

Time frequency analysis was conducted by Short Time Fourier Transform (STFT) via MATLAB R2020a. For task-state data, baseline correction was made using subtraction (Formula 2: *P_BC_*, baseline-corrected power; *P(t,f)*, power value at time-frequency point of (t,f); *P-*(*f*), mean of baseline power values). Regions of interest and time windows were confirmed using point-to-point paired sample *t*-tests of standard and oddball conditions (see S1). Finally, three ROIs corresponding to the P2, N2 and P3 components were identified (P2: 150-220 ms, Fz; N2: 250-320 ms, Cz; P3: 300-600 ms, Oz).

Formula 2:

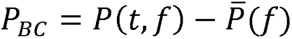

### 2.4 Procedure

All participants were accompanied to the laboratory by their parents. At the beginning, both the children and their parents completed an informed consent form. The children then completed the IVA-CPT and the RSPM. At the same time, the parents were required to complete the three scales mentioned above. After completing these tasks, parents took their child to the shampoo room to have their child’s hair washed. Resting state EEG recordings with eyes open and eyes closed were then collected on all children for three minutes each. Some of the children went on to complete the visual oddball task. Upon completion of all tasks, all children received attention training textbooks and exercise books as gifts.

### 2.5 Data analyses

Statistical analyses were performed using SPSS 25 and MATLAB R2020a. First, Spearman’s correlation analysis was used to screen the collected questionnaire information and EEG information. Each of these variables was correlated separately with the attention and response control scores of the CPT-IVA and significantly correlated variables were included in subsequent general linear models. Notably, neighboring electrodes (that were significant) were averaged to form new variables. Second, stepwise regression was used to build regression models to include objective EEG, ERO data, and subjective parent-rated scores.

## 3 Results

### 3.1 Demographic information

76 participants (30 female, *M_age_* = 8.09, *SD* = 1.81, age range: 6 - 12) completed the resting state EEG acquisition and 46 of them (20 female, *M_age_* = 7.08, *SD* = 1.56, age range: 6 - 12) additionally completed a visual oddball task. For reasons of inability to complete the IVA-CPT and too much EEG noise, 68 eye-closed, 71 eye-open resting-state data, and 41 EEG data under the oddball task were ultimately included in the subsequent data analysis. All participants completed the RSPM and none had intellectual problems.

### 3.2 Variables associated with ADHD symptoms

Spearman’s correlation was used to find parent-rated scores, resting state EEG spectral power, and EROs associated with ADHD attention and response control symptoms. For parental assessments, questionnaire scores commonly used as indicators of ADHD behaviors or tendencies were included in the analysis, including ADHD RS-IV total score, Inattention and Hyperactivity-impulsivity subscale scores, CPRS total score, Factor IV and ADHD Index scores, and CBCL total score. The results showed that only the ADHD RS-IV total score and the Inattention subscale score had a significantly negative correlation with children’s IVA-Attention score (RS-IV total: ρ = -.247, *p* = .038; Inattention: ρ = .292, *p* = .014), indicating that the worse the children’s attention performance, the higher the parents’ scores on the RS-IV total and Inattention scale. All other scale scores were not significant (Table 1).

**Table 1.**
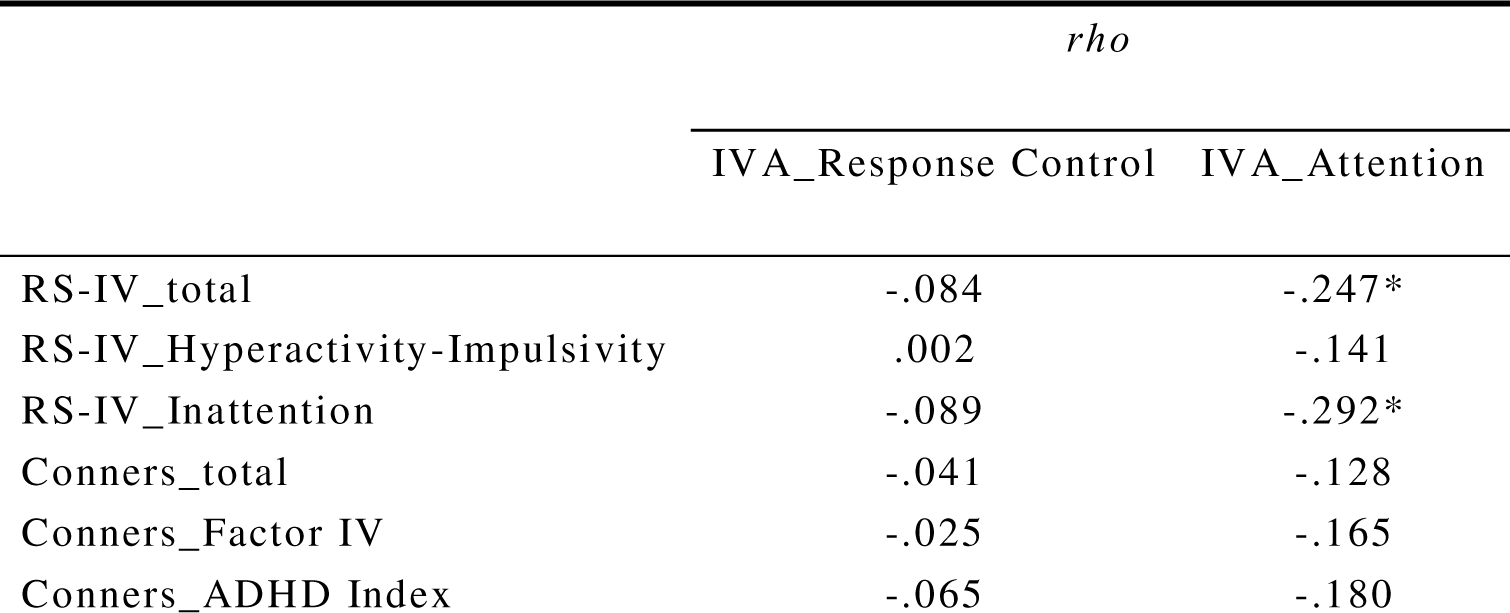
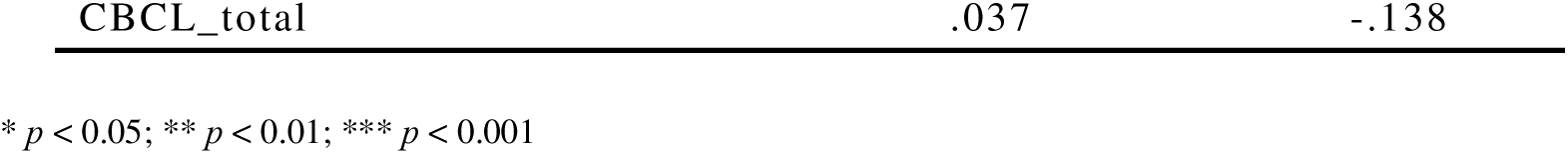
Correlation analysis of parent rating scales and subscales with ADHD symptoms.

Resting EEG band power included the relative power of each electrode in the 4 standard frequency bands (δ, β, α, and θ) in the eyes-open and eyes-closed states. In the eyes-open condition, the delta power of Pz, P1, P3 and P7 positions were negatively correlated with attention performance, and the beta power of P2, POz, PO4, PO6, PO8, Oz and O2 were negatively correlated with response control. In the eyes-closed condition, the beta power of O2 had a significantly negative correlation with children’s attention, with which the theta power of P5, P7, PO3, PO5 and PO7 were positively correlated. In addition, the theta power of P7 and PO3 were positively correlated with response control, whereas the alpha power of AF3, Cz, CP5, CP6, P4, P5, P6, P7, PO3, PO5, PO6, PO7 and PO8 were significantly negatively correlated with it. All significant variables were weakly correlated (.21 < ρ < .40), as shown in Table 2 and Table 3.

**Table 2.**
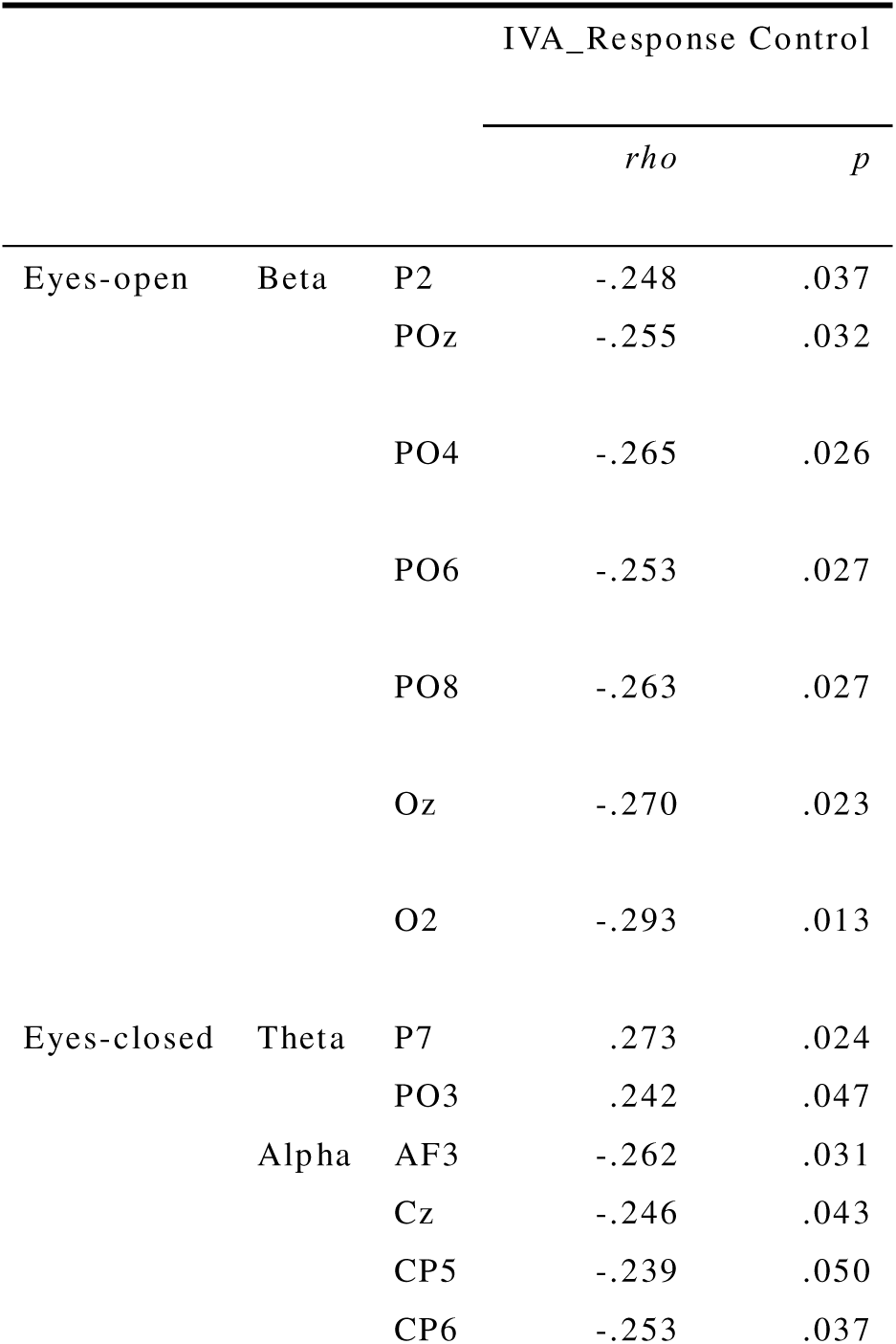
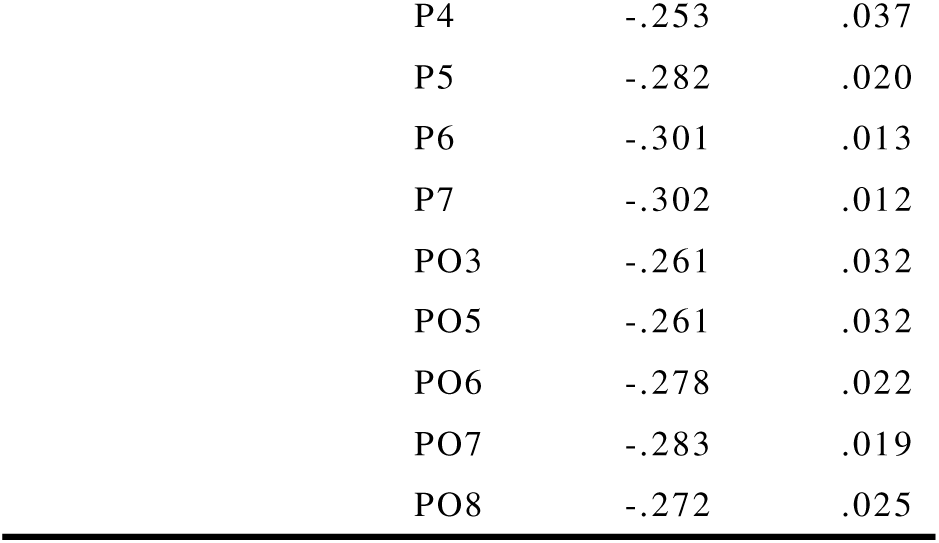
Correlation of relative power across conditions, frequency bands, and electrodes with IVA Response Control score (significant outcomes)

|  |  |  | IVA_Response Control |  |
| --- | --- | --- | --- | --- |
|  |  |  | *rho* | *p* |
| Eyes-open | Beta | P2 | -.248 | .037 |
|  |  | POz | -.255 | .032 |
|  |  | PO4 | -.265 | .026 |
|  |  | PO6 | -.253 | .027 |
|  |  | PO8 | -.263 | .027 |
|  |  | Oz | -.270 | .023 |
|  |  | O2 | -.293 | .013 |
| Eyes-closed | Theta | P7 | .273 | .024 |
|  |  | PO3 | .242 | .047 |
|  | Alpha | AF3 | -.262 | .031 |
|  |  | Cz | -.246 | .043 |
|  |  | CP5 | -.239 | .050 |
|  |  | CP6 | -.253 | .037 |

|  |  |  |
| --- | --- | --- |
| P4 | -.253 | .037 |
| P5 | -.282 | .020 |
| P6 | -.301 | .013 |
| P7 | -.302 | .012 |
| PO3 | -.261 | .032 |
| PO5 | -.261 | .032 |
| PO6 | -.278 | .022 |
| PO7 | -.283 | .019 |
| PO8 | -.272 | .025 |

**Table 3.** Correlation of relative power across conditions, frequency bands, and electrodes with IVA Attention score (significant outcomes)

|  |  |  | IVA_Attention |  |
| --- | --- | --- | --- | --- |
|  |  |  | *rho* | *p* |
| Eyes-open | Delta | Pz | -.242 | .042 |
|  |  | P1 | -.240 | .044 |
|  |  | P3 | -.245 | .040 |
|  |  | P7 | -.236 | .048 |
| Eyes-closed | Beta | O2 | -.249 | .040 |
|  | Theta | P5 | .246 | .043 |
|  |  | P7 | .270 | .026 |
|  |  | PO3 | .265 | .029 |
|  |  | PO5 | .269 | .026 |
|  |  | PO7 | .258 | .033 |

For EROs, the power of three ROIs corresponding to P2, N2, and P3 components was considered (Figure 2). Surprisingly, it was found that no variables were associated with children’s performance on response control. For children’s attention, the P3 component provided the most significant results. The results showed that power in all frequency bands or conditions was significantly negatively correlated with children’s attention, except for delta power in the standard condition. Furthermore, the delta power corresponding to the N2 component of the oddball condition was found to be positively correlated with children’s attention performance (Table 4)

**Figure 2.**
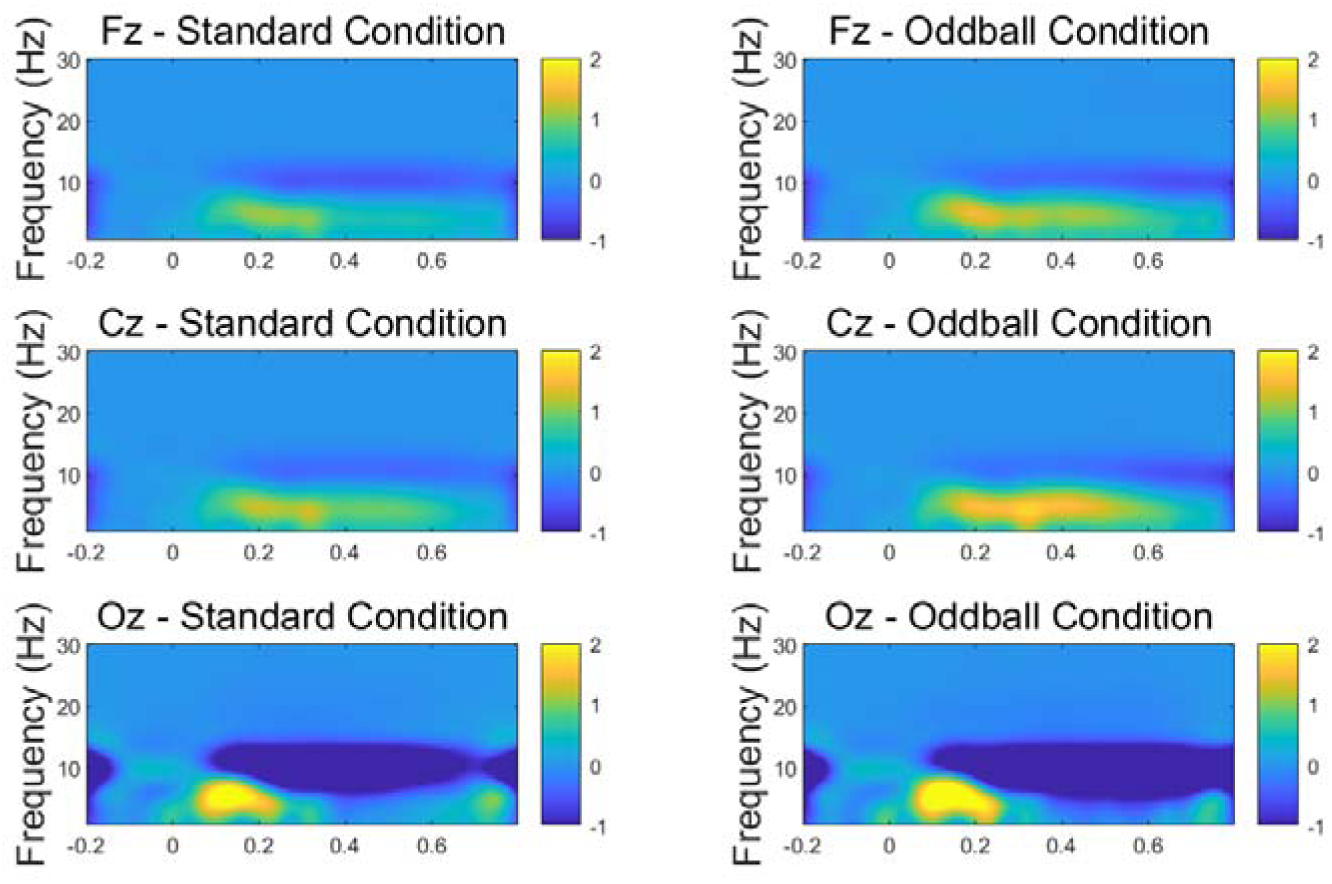
Baseline-corrected time-frequency results of three ROIs corresponding to the P2, N2, and P3 components in the standard and oddball conditions.

**Table 4.** Correlation of the power of three ROIs corresponding to P2, N2, and P3 components with IVA Attention score (significant outcomes)

|  |  |  |  | Power | IVA_Attention |  |
| --- | --- | --- | --- | --- | --- | --- |
|  |  |  |  | *M*±*SD* | *rho* | *p* |
| N2 | Delta | Oddball |  | 2.017±1.610 | .406 | .008 |
| P3 | Delta | Oddball |  | -.301±2.240 | -.321 | .040 |
|  | Theta | Standard |  | -1.463±2.766 | -.310 | .049 |
|  |  | Oddball |  | -2.375±3.999 | -.339 | .030 |
|  | Alpha | Standard |  | -4.125±3.911 | -.414 | .007 |
|  |  | Oddball |  | -5.247±4.777 | -.381 | .014 |
|  | Beta | Standard |  | -.297±0.315 | -.391 | .011 |
|  |  | Oddball |  | -.394±0.316 | -.344 | .028 |

### 3.3 Regression analysis of screened data

#### 3.3.1 Prediction of ADHD symptoms from EEG data

Stepwise regression was used to construct the regression equation, which included variables significantly related to the behavioral performance of children mentioned above as independent variables. IVA attention and control scores, which objectively reflect children’s performance and represent the two core symptoms of ADHD, inattention and hyperactivity/impulsivity, were included as dependent variables.

First, the resting state data of both eyes-open and eyes-closed were added to the stepwise regression. We averaged the power of neighboring significant electrodes to reduce the independent variables and obtained 5 new variables for predicting attention and 12 new variables for response control (see S2). The analysis determined that the relative theta power of the left parietal electrodes and the theta power of the left parieto-occipital electrodes under eyes-closed condition, and the delta power of the middle parietal electrodes with eyes open can predict children’s attention performance separately, but with a minimal adjusted *R^2^* (EC_theta_LP: *B* = 77.234, *SE* = 17.375, *R^2^* = .071, *R_adj_^2^* = .057, *p* = .028; EC_theta_LPO: *B* = 79.554, *SE* = 17.332, *R^2^* = .075, *R_adj_^2^* = .061, *p* = .024; EO_delta_MP: *B* = 114.187, *SE* = 17.447, *R^2^* = .063, *R_adj_^2^* = .049, *p* = .039). Then, the independent variables that contributed the most to the prediction of the dependent variable were added to the model one at a time until we obtained the largest *R^2^* or the prediction was no longer significant. Finally, the best results were obtained when the theta power of the left parieto-occipital electrodes in the eyes-closed condition and the delta power of the middle parietal electrodes in the eyes-open condition were calculated together to predict children’s attention performance (Model 1: *B* = 98.500, *SE* = 16.988, *R^2^*= .125, *R_adj_^2^* = .098, *p* = .013).

For the prediction of response control, it was significant for almost all individual predictor variables, except for the left frontal alpha power in the eyes-closed condition, and the beta power of the middle and right parieto-occipital electrodes in the eyes-open condition (Table 5). As above, the variables were added incrementally, with the best results obtained when both the alpha power of the left parietal and parieto-occipital electrodes under the eyes-closed state were used to predict children’s response control scores (*B* = 119.058, *SE* = 15.802, *R^2^* = .160, *R_adj_^2^* = .134, *p* = .003).

**Table 5.** Prediction of IVA Response Control scores from relative power across conditions, frequency bands, and electrodes.

| Predictor | *B* | *SE* | $R^2$ | $R_{adj}^2$ | *p* |
| --- | --- | --- | --- | --- | --- |
| EC_alpha_LF | 109.217 | 16.718 | .046 | .031 | .080 |
| EC_alpha_C | 112.682 | 16.558 | .064 | .050 | .038 |
| EC_alpha_LCP | 114.687 | 16.373 | .085 | .071 | .016 |
| EC_alpha_LP | 119.381 | 15.857 | .141 | .128 | .002 |
| EC_alpha_LPO | 115.728 | 16.268 | .096 | .083 | .010 |
| EC_alpha_RCP | 116.633 | 16.290 | .094 | .080 | .011 |
| EC_alpha_RP | 118.205 | 16.179 | .106 | .093 | .007 |
| EC_alpha_RPO | 117.984 | 16.247 | .099 | .085 | .009 |
| EC_theta_LP | 84.996 | 16.471 | .074 | .060 | .025 |
| EC_theta_LPO | 89.173 | 16.594 | .060 | .046 | .044 |
| EO_beta_MPO | 109.619 | 16.866 | .029 | .014 | .167 |
| EO_beta_RPO | 110.340 | 16.707 | .047 | .033 | .076 |

EC, eyes-closed; EO, eyes-open; LF, left frontal electrodes; C, central electrodes; LCP, left central parietal electrodes; LP, left parietal electrodes; LPO, left parieto-occipital electrodes; RCP, right central parietal electrodes; RP, right parietal electrodes; RPO, right parieto-occipital electrodes; MPO, middle parieto-occipital electrodes.

Second, as some ERO variables were correlated with children’s attention, we proceeded to conduct a new stepwise regression on the 41 samples that contained task-state data. The results showed that attention scores could not be predicted by any individual resting state EEG variable, but could be predicted by single ERO variables other than the delta power corresponding to P3 in both conditions (Table 6). Furthermore, the stepwise regression analysis demonstrated that the combination of resting state eyes-closed power of right occipital beta and left parieto-occipital theta with alpha power of P3 in the standard condition, beta power of P3 in the oddball condition, and delta power of N2 in the oddball condition exhibited the largest adjusted *R^2^*value for children’s attention performance (Model 2: *B* = 50.814, *SE* = 15.828, *R^2^* = .375, *R_adj_^2^* = .285, *p* = .004).

**Table 6.** Prediction of IVA Attention scores from ERO variables.

| Predictor | *B* | *SE* | $R^2$ | $R_{adj}^2$ | *p* |
| --- | --- | --- | --- | --- | --- |
| N2_delta_oddball | 84.226 | 17.932 | .106 | .083 | .038 |
| P3_delta_oddball | 90.957 | 17.720 | .127 | .104 | .022 |
| P3_theta_standard | 88.471 | 17.821 | .117 | .094 | .029 |
| P3_theta_oddball | 87.775 | 17.640 | .135 | .112 | .018 |
| P3_alpha_standard | 84.004 | 17.400 | .158 | .136 | .010 |
| P3_alpha_oddball | 84.951 | 17.862 | .113 | .090 | .032 |
| P3_beta_standard | 86.633 | 18.117 | .087 | .064 | .061 |
| P3_beta_oddball | 85.172 | 18.170 | .082 | .058 | .070 |

#### 3.3.2 Prediction of ADHD symptoms by combining parental assessments

We added RS-IV scores to the regression model and showed that both total and Inattention subscale scores significantly predicted children’s attention performance (RS-IV_total: *B* = 101.818, *SE* = 17.407, *R^2^* = .067, *R_adj_^2^* = .053, *p* = .033; RS-IV_Inattention: *B* = 104.346, *SE* = 16.979, *R^2^* = .113, *R_adj_^2^* = .100, *p* = .005). Subsequent inclusion of the RS-IV’s inattention score in a stepwise regression showed that it had the largest adjusted *R^2^* in predicting attentional performance in conjunction with the theta power of the left parietal electrodes in the eyes-closed condition and the delta power of the middle parietal electrodes in the eyes-open condition (Model 3: *B* = 103.678, *SE* = 16.286, *R^2^* = .208, *R_adj_ ^2^* = .171, *p* = .002).

The statistical analyses used above were then repeated for the 41 samples that contained task-state data. The results showed that both total and Inattention subscale scores significantly predicted children’s attention performance (RS-IV_total: *B* = 103.645, *SE* = 17.829, *R^2^* = .116, *R_adj_^2^* = .093, *p* = .029; RS-IV_Inattention: *B* = 108.914, *SE* = 16.756, *R^2^* = .219, *R_adj_^2^* = .199, *p* = .002). Stepwise regression showed the best prediction when combining resting state eyes-closed beta power of right occipital electrodes with alpha power of P3 in the standard condition, beta power of P3 in the oddball condition, and delta power of N2 in the oddball condition and parent-rated RS-IV inattention score (Model 4: *B* = 76.556, *SE* = 14.678, *R^2^*= .462, *R_adj_^2^* = .385, *p* < .001). Table 7 displays all models that predict children’s attention performance.

**Table 7.** Comparison of different regression models for predicting IVA attention scores.

| Model | Predictor | | | | | *B* | $R_{adj}^2$ |
| --- | --- | --- | --- | --- | --- | --- | --- |
|  | X1 | X2 | X3 | X4 | X5 |  |  |
| Model 1<br>(*n* = 68) | EC_theta_LPO | EO_delta_MP | - | - | - | 98.500 | .098* |
| Model 2<br>(*n* = 41) | EC_beta_RO | EC_theta_LPO | P3_alpha_standard | P3_beta_oddball | N2_delta_oddball | 50.814 | .285** |
| Model 3<br>(*n* = 68) | EC_theta_LP | EO_delta_MP | RS-IV_Inattention | - | - | 103.678 | .171** |
| Model 4<br>(*n* = 41) | EC_beta_RO | P3_alpha_standard | P3_beta_oddball | N2_delta_oddball | RS-IV_Inattention | 76.556 | .385*** |

EC, eyes-closed; EO, eyes-open; LP, left parietal electrodes; MP, middle parietal electrodes; RO, right occipital electrodes; LPO, left parieto-occipital electrodes.
\* $p < 0.05$ ; \*\* $p < 0.01$ ; \*\*\* $p < 0.001$

## 4 Discussion

The present study aimed to investigate the neural oscillation features of ADHD’s core symptoms, specifically inattention and hyperactivity/impulsivity, in combination with resting state and oddball tasks, and to explore the potential of these characteristics as objective markers of ADHD symptoms. In order to identify the most reliable predictive model, the study also included three parent rating scales. The results of these scales indicated a significant correlation between children’s attention performance and the total and inattention subscale scores of the ADHD RS-IV. This correlation was not observed with other scales or subscales. Furthermore, children’s response control performance was also not found to be significantly correlated with any of the scales or subscales. Power spectrum analysis and time frequency analysis revealed neural oscillation features associated with ADHD symptoms across conditions. Further regression analyses revealed that while certain individual characteristics were predictive of children’s performance, the combination of multiple types of indicators yielded better results. Specifically, the combination of resting state eyes-closed beta power of right occipital electrodes with alpha power of P3 in the standard condition, beta power of P3 in the oddball condition, and delta power of N2 in the oddball condition and parent-rated RS-IV inattention score showed the best prediction.

The following section will discuss these characteristics of both subjective and objective assessments in detail, as well as the potential for applying them to predict ADHD symptoms in children.

### 4.1 Neural oscillation features of ADHD symptoms

Resting state spectral power was collected in four standard frequency bands (δ, β, α, and θ) during both eyes-open and eyes-closed states in children. Our analysis revealed distinct neural oscillation features associated with the core symptoms of ADHD in children. Specifically, we found that increased relative delta activity in the left parietal electrodes during the eyes-open state correlated with poorer attention, while reduced relative theta activity in the left posterior region in the eyes-closed condition was similarly associated with diminished attention performance. In terms of response control function, the eyes-open condition showed that higher beta activity in the right parieto-occipital electrodes was linked to poorer response control. Conversely, in the eyes-closed condition, more active alpha oscillations in the bilateral posterior electrodes and weaker activity in the left parieto-occipital region were indicative of poorer response control performance.

We found that the eyes-open and eyes-closed states exhibited different spectral characteristics. Specifically, the features of theta and alpha bands are prominent in the eyes-closed state, while the beta and delta bands are more characteristic of the eyes-open state. Alpha oscillations occur in the posterior part of the brain and are dominant in the resting, eyes-closed state, playing an important role in human cognition (Bollimunta et al., 2011; Hughes, & Crunelli, 2005). It is generally accepted that the increase in the synchrony of alpha oscillation reflect states of inhibited cortical excitability (Foxe, & Snyder, 2011; Klimesch, 2012). This synchronization not only affects an individual’s detection and perception of external stimuli but is also involved in top-down control for shaping input information and controlling neuronal processing (Doesburg et al., 2016; Wang et al., 2016; Samaha et al., 2015). Our results were consistent with previous findings on the characterization and function of alpha oscillations. We found that when children’s alpha activity was weaker during the resting eyes-closed condition, they exhibited greater control in completing attentional task (i.e., IVA-CPT). Specifically, they performed better at processing irrelevant information and had a stronger ability to inhibit the urge to press. These findings suggest that the emergence of uncontrolled behavior in children may be related to their enhanced alpha activity, indicating that alpha oscillations play a crucial role in children’s behavioral control.

Theta oscillations are another important brain activity in the eyes-closed state that we found to be associated with attentional function in children. Theta oscillations are associated with wide span of cognitive functions (Cavanagh, & Frank, 2014; Korotkova et al., 2018; Soltani-Zangbar et al., 2020). Midfrontal theta activity is thought to be involved in prefrontal control networks, while posterior theta activity is associated with visual attention (Asanowicz et al., 2023; Cavanagh, & Frank, 2014; Guo et al., 2020). Previous studies on ADHD have indicated that children with ADHD exhibit greater theta power compared to typically developing children (Guo et al., 2020; Koehler et al., 2009). Additionally, some research has shown that healthy brain function is closely associated with reduced theta power, suggesting that an impaired ability to respond to external stimuli often correlates with increased theta power (Cao et al., 2022; Klimesch, 1999). However, our findings were inconsistent with previous results and showed that increased theta activation in the parieto-occipital region was linked to better attention performance in children. This may be explained by the age-related changes of theta activity. Some researchers found that during brain maturation in children, theta power decreases with age (Bresnahan et al., 1999; Rodriguez-Martinez et al., 2013), while other studies found that reduced theta activity, which is associated with healthy brain function in children and adults, exhibits the opposite pattern during progressive aging (Cao et al., 2022). These changes may be related to prefrontal control. Greater posterior theta activation might assist individuals in completing attentional processing when the prefrontal lobes are underdeveloped or impaired (Cellier et al., 2021). Overall, the current study found that enhanced theta activity in the posterior part during resting state is a feature of better attentional functioning in children.

Beta and delta oscillations are features of the eyes-open state. The former was found to be negatively related to children’s response control, while the latter was negatively related to children’s attention. Most researches suggested that decreased beta activity is linked to hyperactivity, while some researchers have reported that a small proportion of children with ADHD have excess beta activity and concluded that this pattern of abnormal activity indicated a developmental deviation (Clarke et al., 2013). Clarke et al. (2001a, 2001b, 2013) found that this subtype of children were noted to be more moody and more prone to temper tantrums, but they were not hyperaroused. Although the symptom profile of this group of children was still unclear, both our and previous studies suggest that higher beta activity may be associated with more excessive behavior in children. There has also been some research on delta activity in children with ADHD, and it is generally accepted that elevated frontal relative delta activity is a characteristic of ADHD children, especially in girls (Dupuy et al., 2011, 2013), whereas increased delta activity has been found in comorbid disorders (Clarke et al., 2020). Our results showed that the higher the delta power, the better the children’s attentional functioning, which may reflect the characteristics of our more boys and non-clinical sample.

Overall, there are similarities and differences between the four features we found and previous studies, which suggests that the EEG features associated with ADHD symptoms are influenced by many factors, such as age, gender, and subtype. To verify that our results reflect the characteristics of a particular age group, we explored separate correlation analyses by age and further divided the age groups based on the results and found that some of these features changed at 8-10 years of age (see S3). Evidence from previous EEG research has reported that there is increased theta activity, deficient beta and alpha activation in ADHD children (mainly aged 8-12 years, Clarke et al., 2020), and these activities change with age (Bresnahan et al., 1999; Cellier et al., 2021; Kamida et al., 2016; Rodriguez-Martinez et al., 2013). In contrast, our study recruited children from primary level, included a higher proportion of children aged 6-8 years, and found features mainly in the posterior brain regions. Consequently, the outcomes of this study may be indicative of EEG characteristics associated with attentional functions in younger children with underdeveloped frontal lobes. This evidence suggests that the critical period for these functions may be between the ages of 8 and 10 years.

In addition to resting-state EEG acquisition, the classic visual oddball task was used to record children’s neural oscillation features during attentional processing. We focused on three regions of interest corresponding to the P2, N2, and P3 components, and identified only attentional function-relevant ERO features. Our results mainly showed that the power across all frequency bands and task conditions of P3 component, except for delta power in the standard condition, was significantly negatively correlated with children’s attention. Although we did not find a specific frequency band, these results suggest that 300 to 600 ms after stimulus presentation is a critical period for attentional functioning, and the more active the posterior part of the brain (Oz) is in this time period, the better the child’s attentional performance. These results suggest that late processing of attentional functions may be more complex, recruiting overall brain activity. In addition, we also found a signature of the delta activity in the middle of the brain (about 250-320 ms, Fz) in the oddball condition correlating with children’s attentional function. This result suggests that delta activity may play an important role in early attentional processing.

In ERP studies, P3 (or P300) and N2 components have been well investigated, and it is recognized that the former component is associated with stimulus processing and attentional function, and the latter reflects conflict monitoring and selected attention (Barry et al., 2003; Kok, 2001; Larson et al., 2014). Some studies have considered EROs as a potential mechanism for generating ERPs, where the P300 corresponds to oscillations in the delta and theta frequency bands, and oscillation activity at the N2 latency may be mainly affected by theta activity (Balconi & Pozzoli, 2008). In contrast, our study selected ROIs based on ERP information, but mainly analyzed spectral features corresponding to different stages and brain regions. With time-frequency analysis, the current results may complement the insufficient ERP studies and reflect more the brain activity patterns and underlying mechanisms of attentional processing.

Overall, the EROs information in the visual oddball task may reflect underlying mechanisms at different stages of attentional processing. This information may help to understand children’s brain activation patterns at different attentional stages and with different stimuli. We did not find any ERO feature related to response control, which may be due to the fact that we chose the visual oddball task related to attentional functioning and removed trials in which children responded incorrectly during data processing. In subsequent studies, the classic Go-Nogo task could be included to analyze the brain activity features related to response control in children.

### 4.2 Characteristics and limitations of parental assessment

Three parent rating scales measuring ADHD in children were used in this study. The CBCL and CPRS are commonly used to diagnose ADHD in children and adolescents, while the ADHD RS-IV has good discriminant validity both within the ADHD subtypes and between children with and without ADHD. Our results showed that only the total and inattention subscale scores of the ADHD RS-IV were correlated with children’s attention performance, indicating that the worse the children’s attention performance, the higher the parents’ scores on the inattention dimension.

We found that the three questionnaires performed differently in assessing children’s performance. The parents’ ratings on the CBCL and CPRS scales may not accurately reflect the children’s attention and response control performance. The CBCL is used for assessing a wide range of child emotional and behavioral problems and includes several empirically derived clinical syndrome subscales (Achenbach, 1991; Chang et al., 2016; Gomez et al., 2021). However, because the items included in the Attention Problem dimension varied with age and gender in the normative model of Chinese children, we used only the total score as the outcome of the CBCL (Zhang, 2005). This means that while the CBCL may provide diagnostic information for ADHD assessments, the total score, which offers broadband information, is not as effective in assessing children’s specifical behavioral performance. For the CPRS, although we explored Factor IV (hyperactivity-impulsivity) and the ADHD Index, the results showed that neither the total score nor the subscale scores accurately reflected the children’s true performance. This is consistent with the results of the ADHD RS-IV, which found that parents could accurately assess their children’s attention performance but not their impulsive behavior. These results raise another concern, that parents differ in their identification and assessment of children’s attention and response control performance.

These results confirm our concerns that parents may be more worried about their children’s academic problems, leading to highly subjective ratings of their children’s performance. As our results showed, parents were more sensitive to and accurately rated behaviors related to learning, such as ‘making careless mistakes’, ‘not listening’, and ‘avoiding tasks’. However, they found it difficult to accurately rate behaviors that do not directly affect academic performance, such as ‘running about’, ‘leaving the seat’, and ‘talking excessively’. These findings suggest that diagnoses and treatments based on questionnaires and interviews may be influenced by social context. Although ADHD is a neurodevelopmental disorder characterized by inattention, hyperactivity, and impulsivity, it is also closely linked to parents’ and society’s educational expectations for children (Li, 2024). If a child is not performing well in school, parents are more likely to be reminded by teachers and experience greater academic pressure, leading to increased attention to and even overestimation of the child’s inattentive behaviors. Conversely, if a child is performing well academically but exhibits some hyperactive and impulsive behaviors (this is normal because the academic tasks in the lower grades are very simple), parents may ignore or rationalize these abnormal behaviors.

Overall, in a society where education is highly competitive, parental assessments of the two core symptoms associated with ADHD in children may not fully reflect their child’s objective behavioral performance. This is evidenced by more accurate assessments of inattention and inconsistent assessments of hyperactivity-impulsivity behaviors compared to the children’s actual performance. The results indicate that parental rating scales may not fully capture a child’s true behavioral patterns. There is a pressing need to identify objective indicators, such as EEG biomarkers, to supplement the diagnostic criteria. Additionally, it is important to be cautious of the potential negative consequences of excessive academic stress, which may lead to overassessment and the false belief that diagnosis and medication are necessary to improve academic performance.

### 4.3 Importance of combining multiple indicators

Aiming to provide guidance for clinical diagnosis and classification, we conducted exploratory stepwise regression to compare the predictive abilities of different types of indicators for the two core symptoms of ADHD. Our results show that while some individual characteristics can predict children’s performance, combining multiple metrics offers a more robust prediction, particularly when combining resting-state EEG oscillations, event-related oscillations from the oddball task, and parental ratings. Specifically, including ERO characteristics associated with children’s behavior greatly improved explanatory power compared to using multiple resting-state indicators alone. Additionally, model’s explanatory power further improved with the inclusion of parental assessments.

Previous studies have used EEG features to diagnose ADHD (Lenartowicz, & Loo, 2014; Slater et al., 2022). With the development of machine learning, the use of EEG metrics in diagnostic classification has become increasingly popular, leading to some progress in this field (Ahire et al., 2023; Cao et al., 2023; Deshmukh et al., 2024). Despite of emerging EEG metrics and the application of new technologies, EEG cannot currently be used as a diagnostic tool (Lenartowicz & Loo, 2014). The limited success is perhaps not surprising when considering the heterogeneity in etiology and symptoms in ADHD, as the underlying brain mechanisms are complex.

Currently, the dominant models for explaining ADHD are the hyper- and hypo-arousal models, the maturational lag and the developmental deviation model (Clarke et al., 2020; El-Sayed et al., 2003; Satterfield, & Dawson, 1971; Shaw et al., 2007). Researchers are increasingly recognizing that multi-pathway models should be used to explain ADHD, rather than focusing on single-cause explanations (Lenartowicz, & Loo, 2014; Sonuga-Barke, 2005). This implies that a single EEG metric (such as TBR, frontocentral theta activity, or beta activity) may only predict a specific subtype of ADHD and may not perform well across the general population. In our study, we not only explored a wide range of resting-state eyes-open and eyes-closed spectral information but also examined event-related spectral features of attentional processing that are directly related to attentional function. We included screened EEG features related to children’s performance in stepwise regressions to use as much valid information as possible to jointly predict children’s performance. Our findings confirmed that using multiple indicators to predict children’s behavior yielded better results. Moreover, Arns et al. (2013) noted that heterogeneity is also present in the non-ADHD population. Therefore, we explored using children’s objective behavioral performance as an outcome variable rather than subjective diagnostic information to eliminate the links between the clinical heterogeneity of ADHD and EEG-based metrics. We believe that these results more accurately reflect the underlying mechanisms of normal or abnormal brain’s attention and response control functions.

Overall, our study suggests that combining multiple types of indicators and using children’s objective behavioral performance as a predictor may more accurately and objectively reflect children’s true performance. Furthermore, our exploratory stepwise regression analysis provides novel insights for leveraging big data-based machine learning techniques to identify new indicators for clinical diagnosis.

### 4.4 Contributions and limitations

The current study provides new insights into the EEG characteristics of ADHD symptoms in children, but still has some limitations. First, our study identified EEG features associated with ADHD symptoms and found that these features may change with age and the development of frontal. We propose the conjecture that 8 to 10 years of age may be a critical period for the development of children’s attentional function. Second, our study focused on ADHD symptoms rather than diagnosis, using IVA-CPT scores as children’s objective behavioral performance. We considered that behavior is more directly responsive to the children’s objective symptoms, whereas subjective evaluations from parents may be influenced by academic expectations, social norms, and other factors. Examining symptom-related EEG features might better reflect the underlying nature of the disorder. Furthermore, we propose a novel approach of multi-metric prediction, combining resting-state EEG, task-state EEG, and parental ratings, using two core symptoms of ADHD as outcome variables. On the one hand, we aim to provide insights to address the heterogeneity of ADHD subtypes and non-ADHD populations. On the other hand, our study emphasizes the importance of parental ratings. The combination of EEG indicators is not intended to replace subjective diagnosis but to enhance the accuracy and objectivity of predictions.

There are also some limitations to our study. Due to the difficulty of EEG data acquisition in children, our sample size was relatively small, with only about half of the participants completing the oddball task. Although we found that EEG characteristics may change with age, the current sample was insufficient to support age-specific analyses. Furthermore, we examined EEG features during attentional processing using only one classic visual oddball task. In future studies, increasing the number of participants and incorporating multiple attentional tasks will be essential to validate our results and provide a more comprehensive understanding of the neural activities associated with attentional processing in children.

## 5 Conclusion

In conclusion, the current study analyzed objective and subjective characteristics associated with attentional functioning in children through resting-state EEG acquisition, a visual oddball task, and parent rating scales. We focused on core ADHD symptoms of inattention and hyperactivity/impulsivity rather than diagnosis, using IVA-CPT scores as children’s objective behavioral performance. The main results showed that resting-state eyes-open delta power and eye-closed theta power were associated with children’s attention performance, and eyes-open beta power and eyes-closed alpha power were associated with children’s response control performance. These characteristic activities are predominantly in the posterior part of the brain, suggesting that posterior, rather than frontal, brain activity is closely associated with attentional performance in the early stages of childhood brain development, and 8 to 10 years of age may be a critical period for the development of attentional function. The parent rating scales results showed that only the total and the inattention subscale scores of the RS-IV correlated with children’s attention performance, suggesting that parents may not be able to accurately assess their children’s behavioral performance, especially with regard to behavioral problems that are not related to academics. In addition, we included all variables related to children’s behavioral performance in exploratory stepwise regression analyses and found that the best results were obtained when including all 3 types of assessments, resting-state EEG data, EROs, and parental ratings together. These results provide new insights into multi-metric prediction and some guidance for clinic.

## Data Availability

Data will be made available on request.

## Acknowledgements

We thank Mrs. Jie Song for her invaluable assistance in conducting the experiments.

